# Investigating a structured diagnostic approach for chronic breathlessness in primary care: a mixed-methods feasibility cluster Randomised Controlled Trial

**DOI:** 10.1101/2024.02.21.24303140

**Authors:** Gillian Doe, Jill Clanchy, Simon Wathall, Shaun Barber, Sarah Edwards, Helen Evans, Darren Jackson, Natalie Armstrong, Michael Steiner, Rachael A Evans

## Abstract

**Background:** There is need to reduce delays to diagnosis for chronic breathlessness to improve patient outcomes.

**Objective:** To conduct a mixed-methods feasibility trial of a larger cluster Randomised Controlled Trial (cRCT) investigating a structured symptom-based diagnostic approach versus usual care for chronic breathlessness in primary care

**Methods:** Ten general practitioner (GP) practices were cluster randomised to a structured diagnostic approach for chronic breathlessness, including early investigations (intervention), or usual care. Adults over 40 years old at participating practices were eligible if presenting with chronic breathlessness, without existing diagnosis. The primary feasibility outcomes were participant recruitment and retention rate at one year. Secondary outcomes included number of investigations at three months, and number of diagnoses and patient reported outcome measures (PROMs) at one year.

**Results:** Recruitment rate was 22% (48/220): 65% female, mean (SD) age 66 (11) years, BMI 31.2kg/m^2^ (6.5), median (IQR) MRC dyspnoea 2 (2–3). Retention rate was 85% (41/48). At three months, the intervention group had a median (IQR) of 8 (7–9) investigations compared with 5 (3–6) investigations in usual care. 11/25 (44%) patients in the intervention group had a coded diagnosis for breathlessness at 12 months compared with 6/23 (26%) in usual care. Potential improvements in symptom burden and quality of life were observed in the intervention group.

**Conclusions:** A cRCT investigating a symptom-based diagnostic approach for chronic breathlessness is feasible in primary care showing potential for timely investigations and diagnoses, with PROMs indicating patient-level benefit. A further refined fully powered cRCT with health economic analysis is needed.

**What is already known on this topic:** There are known delays to diagnosis for patients with long-term conditions commonly presenting with breathlessness. A structured symptom-based approach for breathlessness with early investigations may reduce delays and improve patient outcomes, but the clinical and cost effectiveness of such a pathway is unknown.

**What this study adds:** We demonstrated that a future trial investigating a symptom-based structured diagnostic approach for breathlessness is feasible. Our results show participants in the intervention group had more investigations and earlier diagnoses alongside potential to reduce symptom burden.

**How this study might affect research, practice or policy:** A symptom-based approach for breathlessness in primary care has the potential to reduce delays to diagnosis, improve outcomes for patients, and appears acceptable to patients and clinicians; refinement of the pathway and a fully powered cRCT with health economic modelling is needed.

## INTRODUCTION

Breathlessness is a common and distressing symptom with an estimated prevalence of 9-11% in the general population (1, 2), increasing with age to 25% of people over the age of seventy (3). High healthcare use is associated with breathlessness in both primary and secondary care(4–6) and functional impairment from breathlessness is associated with reduced survival (7).

A large proportion of breathlessness is caused by cardiorespiratory disease (8, 9) with clinical data relating to patients over the age of forty indicating the most common causes are Chronic Obstructive Pulmonary Disease (COPD), heart failure (HF), obesity, anaemia and anxiety (9, 10). These conditions may be diagnosed or excluded with investigations frequently available in a primary care setting.

Previous epidemiological studies from primary care have highlighted missed opportunities to diagnose conditions commonly presenting with breathlessness such as COPD and HF, with a large number of patients diagnosed in later stages of the disease or during hospitalisation.(11, 12) Evidence around misdiagnoses for COPD, asthma and Interstitial Lung Disease, (13, 14) also indicates significant challenges in accurate and timely diagnosis for patients. Although there is a National Institute for Health and Care Excellence (NICE) clinical knowledge summary for investigating breathlessness, it does not specify timeframes (15). Recently, using a large UK primary care database (Clinical Practice Research Datalink: CPRD) we have shown that adults diagnosed within six months of presentation with breathlessness have a lower risk of hospital admissions and mortality compared with those waiting beyond six months (16), Breathlessness has also been shown to cause significant burden of ill health among individuals without a confirmed diagnosis (17).

Our overarching hypothesis is that a symptom-based approach for diagnosis in primary care for chronic breathlessness, initiating a holistic suite of diagnostic investigations at the point of presentation, will lead to earlier diagnosis, earlier treatment, and improved outcomes for patients. Early diagnostic investigations for cardiorespiratory conditions leading to earlier holistic treatment may reduce future healthcare contacts and hospitalisations. However, there is clinical equipoise with concerns about over-investigation and over-diagnosis and potential increased associated costs (18).

To investigate the clinical and cost effectiveness of a structured symptom-based diagnostic approach for chronic breathlessness, a large and potentially expensive multi-centre cRCT would be needed. We therefore conducted a feasibility study to inform design of a future trial. The main feasibility aims were:

1. To assess feasibility of participant recruitment and retention rate to enable calculation of the number of GP practices, cluster sizes and duration of the ultimate cRCT.
2. To better understand potential primary outcome measures for the future trial.
3. To understand any influence of the trial design on usual care.

## METHODS

### Study design

We conducted a mixed-methods feasibility study of a multicentre cRCT to investigate a structured diagnostic approach versus usual care for chronic breathlessness in primary care (REC Reference: 19/EM/0201) and registered as a clinical trial (ISRCTN: 14483247).(19) Informed written consent was obtained from all participants.

### Participants

Ten General Practitioner (GP) practices in Leicestershire, UK, were cluster randomised to a structured diagnostic approach (intervention) including early investigations, or usual care. Participants were opportunistically recruited from primary care when they presented to their GP with breathlessness. Eligible participants were over 40 years old, breathless for longer than two months and had not consulted more than twice about their breathlessness. Patients were excluded if they had an existing diagnosis for breathlessness, were acutely unwell requiring hospitalisation, or had an estimated prognosis of less than one year. An electronic template, triggered at the point of consultation by breathlessness Read codes or free text, was used to aid opportunistic recruitment (20).

### Intervention

The structured diagnostic approach included early investigations to be performed within one month (Supplement Figure 1): body mass index (BMI), spirometry, electrocardiogram (ECG), chest X-ray (CXR), Full blood count (FBC), N-terminal (NT)-pro hormone BNP (NT-proBNP) profile, anxiety and depression screening using the Patient Health Questionnaire – 4 item (PHQ-4)(21) and the General Practice Physical Activity Questionnaire (GPPAQ) (22). A diagnostic pathway document was provided for GPs to prompt the investigations and support a structured history and examination (19). In order to ensure participants in the intervention group had all investigations, if they were not performed in primary care, they were completed by the research team where possible.

### Control

The Usual care group were asked to proceed with investigating the patient and their symptoms as per usual practice and were directed to the NICE Clinical Knowledge summary for Breathlessness(15) to standardise care.

### Outcome measures

The primary outcome was recruitment and retention rate to inform future sample and cluster sizes. Recruitment rate per year per practice population size was calculated to provide an estimation of the sample size required for a larger trial. All feasibility measures are listed in Supplement Box 1. Secondary outcome measures included number of investigations at three months, number of diagnoses at 12 months, and time to diagnosis. Patient reported outcome measures (PROMs) were collected at baseline, six and 12 months.

Physical outcome measures were also collected at baseline; collection methods are described in detail in the Supplement (page 2) and protocol paper (19).

PROMs included health-related quality of life (HR-QoL): the Chronic heart questionnaire (CHQ) self-report(23) and EuroQol 5 Dimension 5-Level (EQ5D-5L)(24); breathlessness: Dypsnoea-12(25), Multidimensional Dyspnoea Profile (MDP)(26), Baseline Dyspnoea Index (BDI), Transition Dyspnoea Index (TDI)(27) and the Medical Research Council (MRC) Dyspnoea scale(28); anxiety and depression using the Hospital Anxiety and Depression score(29); patient knowledge and skills to manage their own health using the Patient Activation Measure (PAM).(30) Participants were contacted up to three times for completion and return of their PROM questionnaires.

### Semi-structured interviews

Semi-structured interviews were conducted with patients, clinicians and GP practice staff to understand their experiences of the diagnostic process for breathlessness and taking part in the trial. Interviews were conducted by one of two researchers trained in qualitative methods, recorded and transcribed verbatim. Qualitative data was analysed using thematic analysis (31). Additional feasibility aims were to better understand ‘usual care’ through qualitative analysis of semi-structured interviews with patients and clinicians (reported elsewhere (32)).

### Statistical analysis

Normality of the data was assessed using the Shapiro-Wilkes test. Data are presented as mean (standard deviation [SD]) or median (interquartile range [IQR]). Exploratory data analysis was completed for the secondary outcome measures. The study is not powered to detect statistical differences; the primary feasibility outcome measure was recruitment rate and a sample size calculation was not performed. Recruitment rate is recorded as the proportion of participants consented compared with the number of participants identified as eligible on presentation to their GP. SPSS version 26 was used for statistical analysis. GraphPad Prism 9 was used for all figures presented.

Exploratory analysis was performed on time to diagnosis using survival analyses based on Cox proportional hazards survival modelling. The proportion of patients with valid diagnosis at three months and one year is described and compared using chi squared tests.

## RESULTS

### Recruitment and Retention

Ten out of the fifteen GP practices approached agreed to participate in the study. All practices approached had a patient population of 10,000 or greater, with an Indices of Multiple Deprivation (IMD) quintile range of 1-5. Data available from 7 of the 10 GP practices showed between 6 and 19% of patients identified as eligible from practices were sent to the study team (further detail Supplementary Figure 2).

Recruitment rate was 22% with 48/220 participants recruited between November 2019 and February 2021 (Figure 1): 65% female, mean (SD) age 66 (11) years, BMI 31.2 (6.5), median (IQR) MRC dyspnoea scale grade 2 (2–3) (Table 1). The baseline characteristics between the intervention and usual care groups were similar (Table 1 and 2) except for the ISWT distance, which was higher in the Usual care group, skewed by one individual. The UK COVID pandemic started in March 2020. The recruitment rate before the COVID pandemic ranged from 0.1 to 1 per 1000 GP practice population. Missing data and future cluster sizes calculations are summarised in the Supplement.

**Figure 1.**
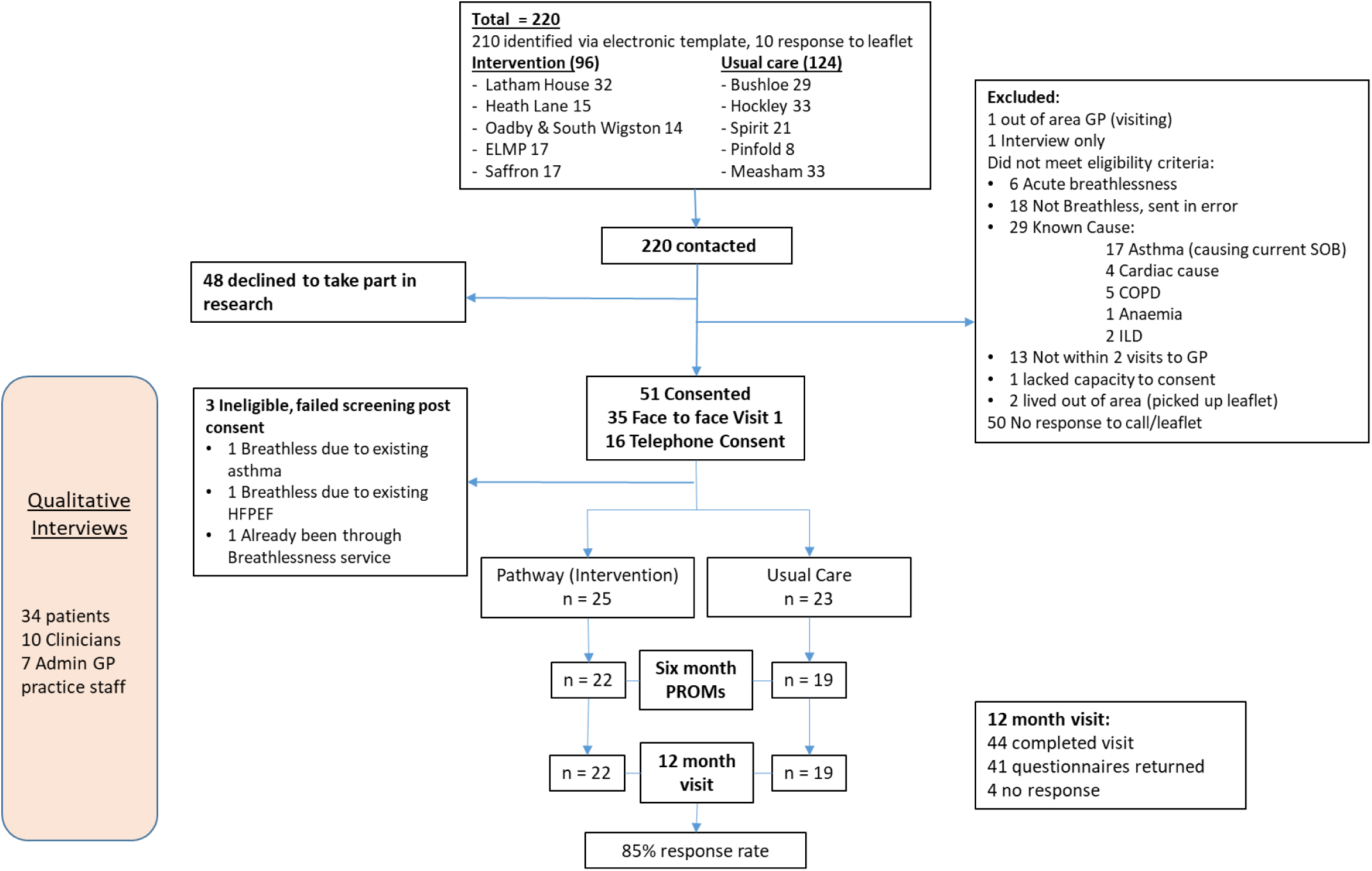
Consort diagram

**Table 1.** Baseline Characteristics.

|  | All participants<br>(n = 48) | Usual Care<br>(n = 23) | Intervention<br>(n = 25) |
| --- | --- | --- | --- |
| Age (years) | 65.8 (11.3) | 64.9 (11.6) | 64.5 (11.3) |
| Gender n (% female) | 31 (65) | 16 (70) | 15 (60) |
| Ethnicity: |  |  |  |
| White | 44 (92) | 21 (92) | 23 (92) |
| Asian/Asian British | 2 (4) | 1 (4) | 1 (4) |
| Black/African/Caribbean/Black British | 2(4) | 1 (4) | 1 (4) |
| BMI (kg/m<sup>2</sup>) | 31.2 (6.5) | 30.8 (6.6) | 31.7 (6.5) |
| IMD (quintile)) | 3 (2-4) | 3 (2-5) | 4 (3-4) |
| MRC Dyspnoea score | 2 (2-3) | 2 (2-3) | 3 (2-3) |
| Smoking status (%) |  |  |  |
| Current | 5 (10) | 2 (9) | 3 (12) |
| Former | 21 (44) | 11 (48) | 10 (40) |
| Never | 22 (46) | 10 (43) | 12 (48) |
| Pack years | 16.0 (5.9-39.2) | 16.0 (6.3-37.5) | 16.0 (5.1-42.0) |
| - Range | 0.2 – 120.0 | 0.2 - 47.0 | 0.25 -120.0 |
| Asbestos exposure - self report (%) | 9 (19) | 7 (30) | 2 (8) |
| Occupational dust exposure - self report (%) | 20 (42) | 10 (44) | 10 (40) |
| Living alone (%) | 14 (29) | 8 (35) | 6 (24) |
| Retired (%) | 28 (58) | 14 (61) | 14 (56) |
| Number of comorbidities | 3 (2-6) | 3 (1-5) | 4 (2-7) |
| Number of medications | 4 (0-5) | 3 (2-6) | 3 (1-6) |
| Rockwood Frailty score: | 4 (3-4) | 4 (3-4) | 4 (3-4) |

Data is presented as Mean (SD), frequency (n, %) or Median (IQR).
*BMI = body mass index, IMD = Indices of Multiple Deprivation, MRC = Medical Research Council Dyspnoea scale. Rockwood Frailty score: 1 - Very fit, 2 – Well no active disease, 3 – Well with treated comorbid disease, 4 – Apparently vulnerable, 5 – Mildly frail, 6 – Moderately frail, 7 – Severely frail.*

**Table 2.** Patient reported outcome measures and Physical outcome measures at baseline.

|  | All participants |  |  | Usual Care |  |  | Intervention |  |  |
| --- | --- | --- | --- | --- | --- | --- | --- | --- | --- |
| PROMs | n = 47 |  |  | n = 22 |  |  | n = 25 |  |  |
| **Dyspnoea -12** | 9.0 (3.0-17.0) |  |  | 8.9 (2.0-13.5) |  |  | 11.9 (4.3-17.5) |  |  |
| **MDP** |  |  |  |  |  |  |  |  |  |
| **A1** | 4.3 (2.4) |  |  | 3.5 (2.3) |  |  | 5.0 (2.4) |  |  |
| **Immediate perception** | 19.0 (13.2) |  |  | 14.9 (12.6) |  |  | 22.5 (13.0) |  |  |
| **Emotional response** | 15.2 (12.4) |  |  | 11.7 (12.3) |  |  | 18.3 (11.7) |  |  |
| **CHQ** |  |  |  |  |  |  |  |  |  |
| **Dyspnoea** | 3.2 (1.2) |  |  | 3.2 (1.1) |  |  | 3.1 (1.3) |  |  |
| **Fatigue** | 3.7 (1.4) |  |  | 3.7 (1.4) |  |  | 3.8 (1.4) |  |  |
| **Emotional Function** | 4.6 (1.3) |  |  | 4.6 (1.4) |  |  | 4.6 (1.3) |  |  |
| **Mastery** | 4.8 (1.4) |  |  | 4.6 (1.3) |  |  | 4.9 (1.6) |  |  |
| **HADS** |  |  |  |  |  |  |  |  |  |
| **Anxiety** | 7.2 (4.9) |  |  | 6.6 (4.9) |  |  | 7.7 (4.9) |  |  |
| **Depression** | 5.6 (3.8) |  |  | 6.1 (4.3) |  |  | 5.3 (3.3) |  |  |
| **EQ5D-5L VAS** | 70.1 (15.8) |  |  | 66.8 (14.6) |  |  | 74.6 (16.2) |  |  |
| **EQ5D-5L Index Value** | 0.77 (0.64-0.85) |  |  | 0.77 (0.67 – 0.85) |  |  | 0.74 (0.43-0.84) |  |  |
| **BDI focal score** | 6.4 (2.1) |  |  | 6.2 (2.3) |  |  | 6.7 (2.0) |  |  |
| **Physical outcomes** | n | % missing |  | n | % missing |  | n | % missing |  |
| **ISWT (m)** | 23 | 52 | 348 (196) | 9 | 61 | 426 (217) | 14 | 44 | 299 (170) |
| **SpO2 post-ISWT (%)** |  |  | 92 (4) |  |  | 93 (4) |  |  | 92 (4) |
| **Peak HR (bpm)** |  |  | 92 (18) |  |  | 103 (17) |  |  | 85 (15) |
| **Peak BORG** |  |  | 3.0 (3.0-4.0) |  |  | 3.5 (2.5-4.0) |  |  | 3.5 (3.0-4.0) |
| **SPPB (score)** | 31 | 35 | 9.0 (7.0-11.0) | 11 | 52 | 8.0 (7.0-11.0) | 20 | 20 | 9.0 (7.0-10.8) |
| **4MGS (seconds)** | 31 | 35 | 4.0 (3.5-5.2) | 11 | 52 | 3.9 (3.6-5.6) | 19 | 24 | 4.1 (3.4-5.1) |
| **Body fat (kg)** | 32 | 33 | 39.4 (9.0) | 12 | 48 | 31.5 (10.0) | 20 | 20 | 35.6 (13.0) |
| **Body fat (%)** |  |  | 34.1 (12.0) |  |  | 38.6 (9.9) |  |  | 39.9 (8.7) |
| **\*Physical Activity:** |  |  |  |  |  |  |  |  |  |
| **Step count** | 32 | 33 | 5011 (2560) | 11 | 52 | 5041 (3090) | 21 | 16 | 4996 (2320) |
| **Sedentary Time (mins)** | 29 | 40 | 660 (113) | 12 | 48 | 649 (93) | 17 | 32 | 668 (128) |

47/48 participants completed the baseline PROMs. Physical outcome measures were collected where possible at face-to-face visits; some participants completed research visits by phone due to the pandemic. The number collected and % missing is presented.
Data is presented as Mean (SD), frequency (%) or Median (IQR). ISWT = Incremental shuttle walk test, SPPB = Short performance physical battery, 4MGS = 4 metre gait speed, CHQ = Chronic Heart Questionnaire (self-report), MDP = Multidimensional Dyspnoea Index, A1 = Affective Domain 1 (relating to breathing discomfort), A2 = Affection domain 2 (relating to emotional responses), PAM = Patient Activation Measure, HADS = Hospital Anxiety and Depression Score, EQ5D-5L = EuroQoL- 5 Dimension 5 level questionnaire, VAS = Visual Analogue Scale, BDI = Baseline Dyspnoea Index (score from 0-12 with a lower score showing worse impairment)
\*Physical Activity: Step count measured via Actigraph waist worn device, Sedentary Time measured via GeneActive device

### Structured Diagnostic Approach versus Usual Care

The Intervention group had a median (IQR) of 8 (7–9) tests compared with 5 (3–6) tests in UC within three months (Figure 2 A). Chest X-ray, blood tests and BMI were the most frequently completed investigations in both Intervention and Usual care groups (Supplement Table 3). Spirometry was unable to be performed for periods of the study due to the COVID pandemic and the reason for non-completion was also recorded (Supplement page 2).

**Figure 2.**
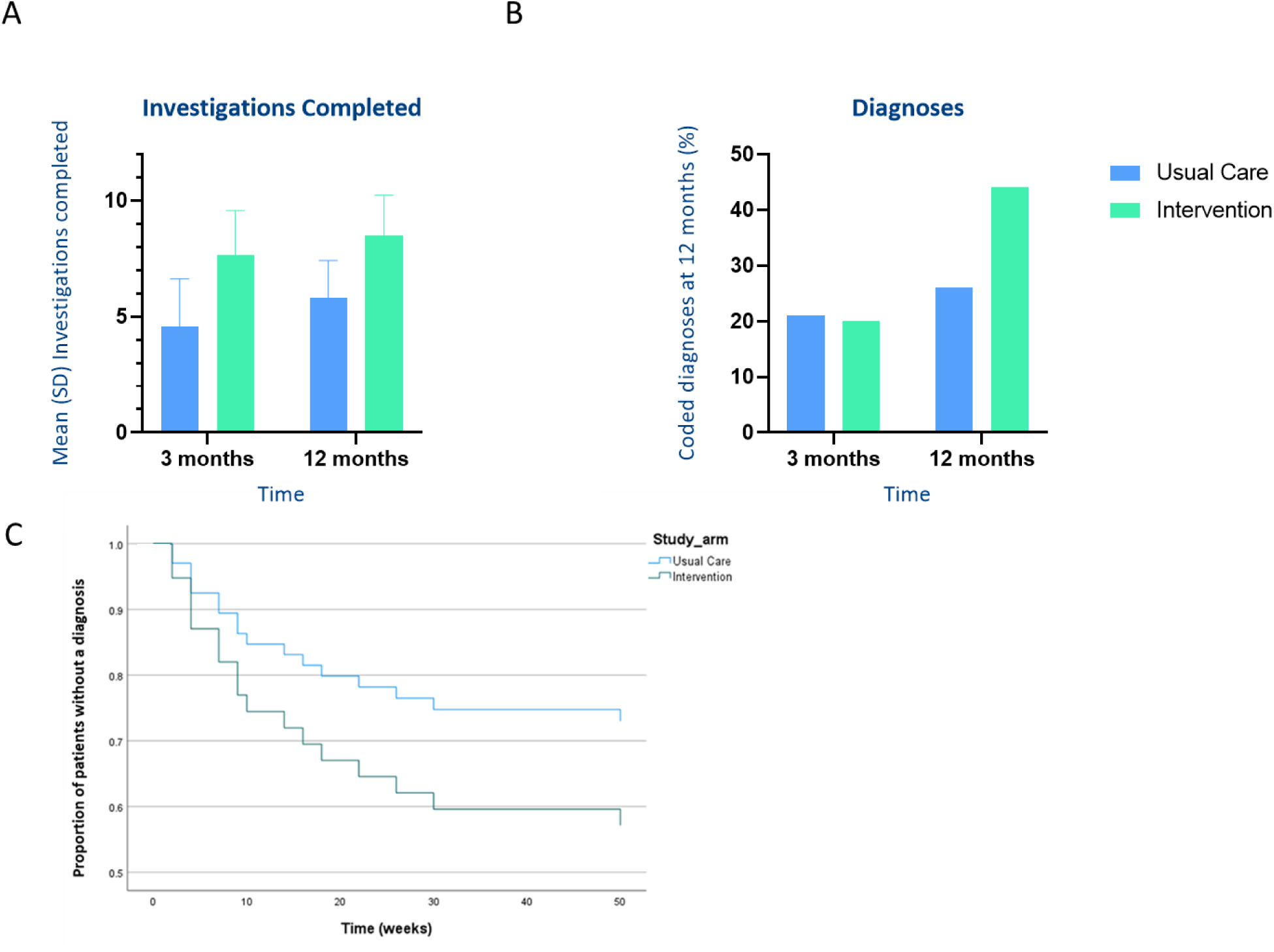
Investigations completed per patient and recorded diagnoses for Intervention group verses Usual care. A. Mean (SD) number of investigations completed per patient at 3 and 12 months. B. Proportion of coded diagnoses for breathlessness at 3 and 12 months. C. Cox proportional hazards survival modelling; proportion of patients without a diagnosis over 1 year.

At 12 months 11 (44%) patients in the Intervention group had a coded diagnoses for their breathlessness versus 6 (26%) patients in usual care (Figure 2 B). Coded diagnoses are summarised in the Supplement. Cox proportional hazards survival modelling (Figure 2 C) derived a non-significant hazard ratio of 1.78 (95% CI 0.7-4.8, p=0.26) indicating the Intervention group had 78% greater chance of diagnosis relative to the Usual care group. The proportion of patients with valid diagnosis was compared between groups at three months; *X^2^* (1, n = 48) = 0.02 p=.88, and 12 months; *X^2^* (1, n = 48) = 1.68 p=.20.

For the PROMs, the mean difference between the intervention and usual care groups from baseline to 12 months was greater than the minimal clinical important difference (MCID) for symptom burden and quality of life: the MDP immediate perception (-15.4 ± 3.5) and emotional response (-8.5 ± 3.8) domains, the Dyspnoea 12 (-6.3 ± 2.6), the Dyspnoea domain of the CHQ (1.0 ± 0.5) and the Utility Index for the EQ5D-5L (0.12 ± 0.07) (Table 3 and Supplement Figure 3). Symptom burden as measured by PROM questionnaires appeared higher in the intervention group at baseline. No improvement was seen in the Hospital Anxiety and Depression score.

**Table 3.** Comparison of Patient Reported Outcomes Measures (PROMs) for 6 and 12 month responders.

|  | *Usual care (n=19)* |  |  | *Intervention (n=22)* |  |  |  |  |
| --- | --- | --- | --- | --- | --- | --- | --- | --- |
|  | **Baseline** | **6 months** | **12 Months** | **Baseline** | **6 months** | **12 Months** | **MCID** | **Mean group difference (IG-UC) change from baseline at 12 months** |
| **MDP<sup>†</sup>** |  |  |  |  |  |  |  |  |
| *Immediate perception* | 14.0 (12.9) | 13.7 (14.0) | 16.6 (15.2) | 24.8 (12.1) | 13.6 (10.6) | 12.0 (10.1) | - 4.6 | -15.4 (3.5)* |
| *Emotional response* | 9.8 (12.2) | 8.4 (12.0) | 10.9 (12.5) | 18.6(12.0) | 11.9(11.9) | 11.7 (13.1) | - 2.4 | -8.5 (3.8)* |
| **Dyspnoea-12<sup>†</sup>** | 7.4 (5.8) | 9.4 (7.9) | 9.1 (9.0) | 12.7 (8.2) | 8.7 (8.4) | 8.1 (7.0) | - 2.8 | -6.3 (2.6)* |
| **Nijmegen** | 16.3 (8.4) | 17.8 (10.8) | 15.6 (9.4) | 22.2 (10.1) | 19.2 (10.3) | 17.5 (11.1) | N/A | -5.1 (2.6) |
| *% with Score indicating HVS* | 26 | 26 | 26 | 41 | 27 | 30 |  |  |
| **CHQ** |  |  |  |  |  |  |  |  |
| *Dyspnoea* | 3.3 (1.2) | 3.6 (1.4) | 3.7 (1.4) | 3.0 (1.3) | 4.5 (1.4) | 4.3 (1.7) | +0.5 | 1.0 (0.5)* |
| *Fatigue* | 4.0 (1.3) | 3.8 (1.4) | 4.0 (1.3) | 3.7 (1.5) | 4.1 (1.4) | 4.0 (1.4) | +0.5 | 0.3 (0.3) |
| *Emotional Function* | 4.9 (1.4) | 5.0 (1.4) | 5.0 (1.4) | 4.6 (1.2) | 5.0 (1.3) | 4.7 (1.5) | +0.5 | 0.1 (0.3) |
| *Mastery* | 4.9 (1.1) | 5.0 (1.5) | 5.1 (1.5) | 4.8 (1.6) | 5.1 (1.3) | 5.2 (1.5) | +0.5 | 0.2 (0.5) |
| **HADS** |  |  |  |  |  |  |  |  |
| *Anxiety* | 5.7 (4.1) | 5.6 (4.6) | 5.3 (4.3) | 7.5 (4.5) | 5.7 (3.1) | 7.3 (4.5) | - 1.7 | 0.3 (1.0) |
| *Depression* | 5.6 (3.7) | 5.3 (4.0) | 6.1 (4.2) | 5.3 (3.2) | 4.7 (4.4) | 6.0 (4.9) | - 1.7 | 0.3 (1.0) |
| **EQ5D-5L** |  |  |  |  |  |  |  |  |
| *Index Score* | 0.76 (0.16) | 0.70 (0.33) | 0.72 (0.25) | 0.63 (0.31) | 0.76 (0.20) | 0.71(0.26) | + 0.051 | 0.12 (0.07)* |
| *VAS* | 68 (15) | 66.3 (18.2) | 67 (20) | 74 (17) | 74.30 (14.8) | 67 (19) | + 6.9 | -6 (5) |
| **Patient Activation Measure** | 55.5 (10.9) | 56.5 (12.9) | 55.2 (9.3) | 59.2 (14.7) | 61.2 (15.6) | 56.9 (15.5) | +4.0 | -1.2 (2.9) |

MDP = Multidimensional Dyspnoea Profile, HVS = hyperventilation syndrome, CHQ = Chronic Heart Questionnaire, HADS = Hospital Anxiety and depression Score, EQ5D-5L = EuroQol 5 Dimension 5 Level questionnaire, VAS = Visual Analogue Scale. <sup>†</sup>MDP and Dyspnoea-12: reduction in score = improvement. \*Mean group difference (IG-UC) from baseline to 12 months greater than MCID

### Understanding Usual Care and influence of the trial

Thirty-four patient participants, ten clinician participants, and seven GP practice staff completed semi-structured interviews. Patients: 20 (59%) were female, mean (SD, range) age 68 (10.8, 48-89) years, 32 (94%) white British, 1 Black African and 1 Asian British, median (IQR) indices of multiple deprivation quintile 3 (2–5). The clinicians had mean (SD, range) of 17 (6.3, 6-30) years’ experience, five (50%) were female, 3 were Asian/Asian British and 7 were white British, 9 were GPs and 1 respiratory Nurse. Six (86%) of the GP practice staff were female and all were white British. The trial experience qualitative data are presented in Table 4.

**Table 4.** Feasibility measures collected from interviews with patients and clinicians.

|  |  |
| --- | --- |
| **Time for GPs to screen for eligibility using the electronic template pop up**<br>(Clinician and GP practice admin staff quotes) | *“Well, I have to say it’s been very unobtrusive hasn’t it. Because all that you’ve been asking us to do is ask the patient.”* (Clinician)<br>*“I think on SystmOne as soon as you type breathlessness all of the information comes up which is really great. I think it prompts people to think about the study and to think about, is this patient possibly suitable?”* (Clinician)<br>*“There’s a few GPs that get irritated by too many pop-ups, so I’ve had the odd comment about it. But I think that’s sometimes more a reflection of just the general stress and tiredness that everyone’s feeling at the moment more than anything.”* (Practice Staff)<br>*“But yeah when it popped up, not a problem... so it’s a fairly straightforward would you be interested or not?”* (Clinician)<br>*“...since COVID, everybody’s breathless, so it popped up more times than it probably should have, because obviously more people are becoming more breathless with COVID and things like that. But before that I think it worked pretty well, because it’s just like a little reminder to the GPs to ask if they want to participate.”* (Practice Staff) |
| **Acceptability of the research visit to patients**<br>(Patient quotes) | *“I was very interested in it and I was very pleased to do the exercises and that to see, so that somebody else could see how good or bad I was, you know, with my breathing and that.”* (Patient)<br>*“It was all right once I was there and I did the tests, it was all right once I got back. It was a long day though”* (Patient)<br>*“Because you’ve taken time to explain things. Because there was a lot of good clear information sent out at the start.”* (Patient)<br>*“I had loads of forms, and I’m dyslexic, you helped me all through that though.”* (Patient) |
| **Participant experience of the trial**<br>(Patient quotes) | *"I found it helpful, probably found it helpful just to talk as well, you know, to be able to talk to somebody about it [breathlessness]; instead of just, I suppose instead of just worrying about it, you know."* (Patient)<br>*"I think it's been quite good. And I think it helps people to offload a bit as well, and think that somebody's taking notice. I think that's really important. To think that somebody's actually going and researching and trying to make a difference, that's important, especially if it's like me when they think doctors aren't listening, and thinking how important it is and how much it's affecting people's lives."* (Patient)<br>*"Well I think from my perspective lovely, because somebody's interested in what I'm doing. But as far as the study goes, I've not found it intrusive or difficult... made to feel as though they're valued and important. So yeah I think it's a good thing. I'm interested in what you're doing."* (Patient)<br>*"...you can talk about things that you wouldn't normally talk about, to be fair, I mean, I wouldn't say what I've just said to you to anybody else, to anybody, because the doctors don't want to know that, understandable but no."* (Patient) |
| **GP experience of participating in the trial and influence on their practice**<br>(Clinician quotes) | *"...the thought crossed my mind as to whether or not if I would do things differently. But no I don't think it has because I think I would still do what I think is right for the patient... I was fairly confident that how we manage things in the practice I think we practice a good level of medicine, so I think I don't mind the fact that we weren't put into the intervention trial. So, I think that didn't bother me really."* (Clinician).<br>*"From my point of view, I would possibly say no [to influence on practice], but that's just myself, because I don't think there's anything really that I wouldn't have already done in terms of investigation, how I've managed these patients. I suppose the only, thinking about it is I know in your study you've got the questionnaires haven't you, the more mental health side questionnaires. I suppose whether that side of it, I'm more, I guess maybe more aware of that being a potential source of patient symptoms, the anxiety side of things."* (Clinician)<br>*I think that does make you think about what you're doing more. I mean from a, you know, you try not to change what you're doing but I suppose you're a little bit more cautious... you probably make a little bit more of an effort to write things more clearly and be a bit more thorough.* (Clinician)<br>*It's perhaps just thinking about how we teach GPs to approach it generally and about how we code breathlessness and what approach we take.* (Clinician) |

All participants interviewed reported that taking part in the study was a positive experience. Participants commented there were a lot of questionnaires to complete, some of which were difficult to understand. One participant had additional needs to complete the questionnaires and the researcher supported them to ensure the questionnaires were still answered in a ‘self-report’ way.

Clinicians and practice staff were mostly satisfied with the experience of being in the study. Views about using the electronic template for opportunistic participant recruitment were largely positive, in particular the low burden on time in a consultation, and are described in more detail elsewhere.(20) The role of the GP practice to recruit patients appeared to be acceptable and interviewees expressed that although overall they did not feel being in the trial influenced their practice, it made them more aware of the contributing factors to breathlessness (such as anxiety) and the need to be clear in their documentation.

## DISCUSSION

Our overarching aim is to improve the symptoms, quality of life and survival for adults living with chronic breathlessness through earlier diagnosis using an affordable approach for the healthcare system. Through a feasibility study, we show that a future cRCT investigating a structured diagnostic approach for breathlessness is feasible in primary care, demonstrated by a 22% recruitment and 85% retention rate. Our results indicate that the proposed symptom-based investigative approach, with parallel completion of early investigations rather than the usual incremental approach (32, 33), supports the potential to improve time to investigations and diagnoses for patients. The patient reported outcomes indicated potential patient level benefit with this approach including symptoms and quality adjusted life years (EQ5D-5L at one year).

Delays to diagnosis for people presenting with breathlessness are well documented and we have recently shown in a cohort study that 33% of 101,369 patients did not receive any diagnosis within two years of presentation with breathlessness (16). We also reported that delays to diagnosis was associated with higher risk of hospital admissions and mortality in the subsequent two years (16). Our symptom-based approach used in the current study contrasts with current disease-specific clinical algorithms for assessment and diagnosis described in a review (34) whereby a stepwise approach is utilised for investigation and in a ‘disease silo’ from other potential contributing conditions. Many of the studies identified used patient history, physical examination, full blood count, chest X-ray and ECG as the first stage in their diagnostic algorithm but without a specified timeframe. Importantly only one study in the review was undertaken in primary care, highlighting the lack of evidence despite primary care being the most likely place of first presentation with breathlessness as cross sectional data suggest breathlessness accounts for 4% of consultations in primary care (5).

National Health Service England (NHSE) have developed and recently published a diagnostic tool for breathlessness (33) which closely aligns with the diagnostic investigations utilised in the current study, but did not advocate performing an early panel of investigations, rather it provides flexibility to complete initial investigations according to clinical judgement.

However, our data from CPRD highlights the current delays to diagnosis and associated worse outcomes from the latter approach. Our previous research highlighted the possible reasons for delay to diagnosis included challenges with symptom recognition, timely investigations and confirming a positive diagnosis (32). An incremental approach to investigation to rule out individual diagnoses was described by clinicians, which aligns with disease-specific guidelines which promote excluding a particular diagnosis, rather than a holistic approach to find all causes of a symptom. Following an incremental approach could be appropriate if timely investigations and multiple reviews were possible; however, this is commonly not achievable, would use more clinician time, and has been further exacerbated by the COVID-19 pandemic causing delays in healthcare (35). A symptom-based approach also enables identification of multiple causes of breathlessness which is important and relevant as multi-morbidity prevalence rises and is a major problem for healthcare systems (36). There remains clinician equipoise between using an early parallel investigations approach versus sequential investigations, but our study supports the former and a larger trial is feasible.

Our research also raises the question of which other investigations could be included in a diagnostic pathway in primary care with a desirable criterion of being low cost, accessible, sensitive and specific. We found that a holistic approach to breathlessness was often absent and screening for anxiety and depression was not routinely recorded as part of usual care with a marked difference between the groups; 84% in the Intervention group, compared with 8% in usual care. Even in the intervention group, anxiety and depression screening was frequently picked up at the research visit having not been completed in primary care. A common screening tool is the four item PHQ-4 screening tool and this can be routinely embedded in electronic patient healthcare systems. Given the high prevalence of anxiety and depression associated with breathlessness (9), it is importance to include screening as part of the diagnostic approach in the breathless patient population (37). We also need to consider the impact of breathing pattern disorder (BPD) as a cause for breathlessness and a future diagnostic approach may need to include assessment for BPD. Tools to assess BPD include the Breathing Pattern Assessment Tool (BPAT) and the Nijmegen questionnaire can screen for hyperventilation, but neither are commonly used in primary care and BPD diagnosis often requires clinician expertise. Breathing pattern disorder has become more commonly seen in primary care due to Long Covid (38).

Asthma was the most common diagnosis in both groups of the current feasibility study despite the population being over 40 years old. We purposefully chose not to include Fractional exhaled Nitrous Oxide (FeNO) in our panel of investigations due to the diagnostic approach of having all investigations for all patients in that cluster. FeNO is a relatively quick and easy investigation to complete in primary care, with NO as a biomarker of type-2 airway inflammation commonly seen in the diagnosis of asthma (39). However, as with all tests it needs a high pre-test probability of asthma to be helpful. We made an assumption that the population over 40 were at high risk of the common conditions our panel were able to either diagnose or exclude and tested whether doing all the investigations as a panel led to further diagnoses. Further refining the approach to add individual risk stratification for chronic cardiorespiratory disease might help reduce any unnecessary investigations.

Similarly, increasing the complexity of the pathway to include assessment of the pre-test probability of asthma would help to suggest how FeNO testing should be integrated. We only tested rather basic investigations in the current study, but still showed the potential for a positive signal for the majority of outcomes. Research is ongoing to understand the risk factors for breathlessness using machine learning that could also be added to a future algorithm (40).

Advances are being made with biomarkers other than blood tests; the role of exhaled breath volatile organic compounds in differentiating acute breathlessness has been explored as an option for non-invasive diagnostics in acute settings with cardiorespiratory patients(41). It is not currently known how this could translate into primary care but there is urgent need for novel diagnostics particularly for airways disease.

Whilst we focused on performing simple, basic investigations which should be readily accessible in primary care (notwithstanding the challenges with spirometry (42)), we acknowledge this is only the first step in the diagnostic process. However, even by influencing the investigations at this early step, 44% of patients in the intervention group of the cRCT had a coded diagnosis for their breathlessness at 12 months compared with 26% in UC. These investigations (and the time saved by doing them early and in parallel) should help in selecting further investigations and/or specialist review. A further feature of our approach locally is a joint cardiorespiratory specialist clinic for unexplained breathlessness after the panel of investigations. Patients in both clusters could have been referred to this clinic by their GP and therefore reduced the effect of the intervention. We only used a ‘coded diagnosis’ to reflect the healthcare record used by clinicians and to identify further healthcare including vaccination schedules and review appointments for example. There were further diagnoses made that could be seen in the free text of appointments that were not coded.

We have previously reported that breathlessness management is an unmet need whilst awaiting a diagnosis and others report the wider unmet need (4). Although this was a feasibility study symptom burden and quality of life outcomes in our study indicate possible patient benefit for those in the intervention group, but we acknowledge the importance and necessity of specific breathlessness self-management and therapies including exercise rehabilitation in addition to the diagnosis and disease specific treatment.

### Strengths and limitations of the Study

Due to the study recruitment period, it is anticipated that the pandemic and the subsequent impact on primary care processes may have reduced the number of patients presenting to their GP (35, 43), willingness to participate in the study, and availability of some of the diagnostic tests particularly spirometry. Spirometry was halted entirely in primary care from 2020, during our study period (44). The recruitment rate prior to the COVID pandemic is likely more relevant for future trial design. Our study employed a pragmatic approach with the intervention embedded in clinical care at the GP practice level, opportunistic recruitment at point of patient presentation, and adaptation to the design allowing continued recruitment through the COVID pandemic. Opportunistic recruitment was a successful approach in this study design to identify a patient by a symptom at presentation and has been shown to be of benefit in previous primary care studies. There are many identified barriers to recruiting to research in primary care, including insufficient funding, resource and research experience in GP settings (45). Our work has demonstrated that signposting patients about research at the point of presentation to healthcare, while reducing the burden on clinicians to discuss the research in detail, is a helpful approach (20).

Cluster randomisation at the level of the GP practice was selected to reduce the risk of contamination in usual care and this appeared successful. The proposed diagnostic tool and future trial design might require further refinement. Most participants recruited were of white British ethnicity which is not representative of the diverse ethnic backgrounds of our local population. Provision of translation services would be required for a future trial with engagement with diverse patient and public representation embedded in the trial design to improve this (46).

## CONCLUSION

Our results indicate that a cRCT investigating a structured diagnostic approach for chronic breathlessness is feasible in primary care. Improving patient care and experience for those living with breathlessness requires prompt and accurate diagnosis, allowing access to appropriate treatment and support. The structured diagnostic approach for chronic breathlessness used here appeared acceptable to patients and clinicians, with the potential to achieve more timely investigations and explanatory coded diagnoses, leading to potential patient level benefit at six and 12 months. We report a positive indication that early investigation as part of a structured diagnostic approach is of benefit but further refinement and a fully powered cRCT with health economic analysis would be needed to evaluate the clinical and cost effectiveness.

## Supporting information

Supplement Information feasibility cRCT Breathe DEEP

## Acknowledgements

We acknowledge and are grateful to our public and patient involvement (PPI) members; Winifred Smart, Jagruti Lalseta, Brian Davies, Paul Ashby and Tony Watling. RAE thanks Professor Mike Morgan and the NIHR Research Design Service East Midlands, University of Leicester for their support with the NIHR clinician scientist fellowship application. Thank you to Stacey Chantrell for her support with the qualitative interviews and Dr Noel Baxter for his advice and expertise with the diagnostic pathway design. We also thank the clinicians who contributed to the development of the structured diagnostic pathway including Professor Ruth Green, Mrs Louise Clayton, Mrs Karen Moore, Mr Alex Woodward, Mrs Jo Szymkowiak, Ms Alison Scott, Ms Jane Giles. The work is supported by the NIHR Leicester Biomedical Research Centre-Respiratory.

## Contributors

RAE conceived the research idea, and developed the theory and plan for the study with MS and DJ. GD drafted the initial manuscript amended by RAE. SW developed the electronic template for use in the recruitment strategy. All authors (RAE, GD, JC, SW, SE, SB, HE, DJ, NA and MS) contributed to the study development and reviewed, commented and approved the manuscript.

## Patient consent for publication

Written informed consent was obtained from all patients.

## Ethics approval

provided by the Nottingham 1 Research Ethics Committee (REC Reference: 19/EM/0201) and registered as a clinical trial (ISRCTN: 14483247) (19). Informed written consent was obtained from all participants.

## Data availability statement

Data are available on reasonable request from the corresponding author.

## Supplemental material

The supplemental material has been provided by the authors.

## REFERENCES

1. Currow DC, Plummer JL, Crockett A, Abernethy AP. A community population survey of prevalence and severity of dyspnea in adults. J Pain Symptom Manage. 2009;38(4):533–45.

2. Bowden JA, To TH, Abernethy AP, Currow DC. Predictors of chronic breathlessness: a large population study. BMC Public Health. 2011;11:33.

3. Smith AK, Currow DC, Abernethy AP, Johnson MJ, Miao Y, Boscardin WJ, et al. Prevalence and Outcomes of Breathlessness in Older Adults: A National Population Study. J Am Geriatr Soc. 2016;64(10):2035–41.

4. Hutchinson A, Pickering A, Williams P, Bland JM, Johnson MJ. Breathlessness and presentation to the emergency department: a survey and clinical record review. BMC Pulm Med. 2017;17(1):53.

5. Frese T, Sobeck C, Herrmann K, Sandholzer H. Dyspnea as the reason for encounter in general practice. J Clin Med Res. 2011;3(5):239–46.

6. Stevens JP, Dechen T, Schwartzstein R, O’Donnell C, Baker K, Howell MD, et al. Prevalence of Dyspnea Among Hospitalized Patients at the Time of Admission. Journal of pain and symptom management. 2018;56(1):15–22.e2.

7. Frostad A, Soyseth V, Andersen A, Gulsvik A. Respiratory symptoms as predictors of all-cause mortality in an urban community: a 30-year follow-up. J Intern Med. 2006;259(5):520–9.

8. Chen Y, Hayward R, Chew-Graham CA, Hubbard R, Croft P, Sims K, et al. Prognostic value of first-recorded breathlessness for future chronic respiratory and heart disease: a cohort study using a UK national primary care database. Br J Gen Pract. 2020;70(693):e264–e73.

9. Sandberg J, Ekstrom M, Borjesson M, Bergstrom G, Rosengren A, Angeras O, et al. Underlying contributing conditions to breathlessness among middle-aged individuals in the general population: a cross-sectional study. BMJ Open Respir Res. 2020;7(1).

10. IMPRESS. Breathlessness IMPRESS Tips (BITs) For clinicians 2016 [Available from: https://www.researchgate.net/publication/260362662_Breathlessness_IMPRESS_Tips_BITs_for_clin icians.

11. Jones RC, Price D, Ryan D, Sims EJ, von Ziegenweidt J, Mascarenhas L, et al. Opportunities to diagnose chronic obstructive pulmonary disease in routine care in the UK: a retrospective study of a clinical cohort. Lancet Respir Med. 2014;2(4):267–76.

12. Bottle A, Kim D, Aylin P, Cowie MR, Majeed A, Hayhoe B. Routes to diagnosis of heart failure: observational study using linked data in England. Heart. 2018;104(7):600–5.

13. Heffler E, Crimi C, Mancuso S, Campisi R, Puggioni F, Brussino L, et al. Misdiagnosis of asthma and COPD and underuse of spirometry in primary care unselected patients. Respir Med. 2018;142:48–52.

14. du Bois RM. An earlier and more confident diagnosis of idiopathic pulmonary fibrosis. Eur Respir Rev. 2012;21(124):141–6.

15. NICE. Clinical Knowledge Summary on Breathlessness 2017 [Available from: https://cks.nice.org.uk/breathlessness.

16. Karsanji U, Lawson C, Bottle A, Doe G, Khunti K, Quint J, et al. Time to diagnosis and long-term outcomes for adults presenting with breathlessness. medRxiv 2024 doi: 10.1101/2024.02.19.243026180.

17. Poulos LM, Ampon RD, Currow DC, Marks GB, Toelle BG, Reddel HK. Prevalence and burden of breathlessness in Australian adults: The National Breathlessness Survey-a cross-sectional web-based population survey. Respirology. 2021;26(8):768–75.

18. Singh H, Dickinson JA, Theriault G, Grad R, Groulx S, Wilson BJ, et al. Overdiagnosis: causes and consequences in primary health care. Can Fam Physician. 2018;64(9):654–9.

19. Doe G, Clanchy J, Wathall S, Chantrell S, Edwards S, Baxter N, et al. Feasibility study of a multicentre cluster randomised control trial to investigate the clinical and cost-effectiveness of a structured diagnostic pathway in primary care for chronic breathlessness: protocol paper. BMJ Open. 2021;11(11):e057362.

20. Doe G, Wathall S, Clanchy J, Edwards S, Evans H, Steiner MC, et al. Comparing research recruitment strategies to prospectively identify patients presenting with breathlessness in primary care. NPJ Prim Care Respir Med. 2022;32(1):49.

21. Kroenke K, Spitzer RL, Williams JB, Löwe B. An ultra-brief screening scale for anxiety and depression: the PHQ-4. Psychosomatics. 2009;50(6):613–21.

22. DepartmentofHealthandSocialCare. General Practice Physical Activity Questionnaire. 2013.

23. Evans RA, Singh SJ, Williams JE, Morgan MD. The development of a self-reported version of the chronic heart questionnaire. J Cardiopulm Rehabil Prev. 2011;31(6):365–72.

24. (5Q5D5L). E. Euroqol (5Q5D-5L) 2016 [Available from: www.euroqol.org/eq-5d-products/eq-5d-5l.html.

25. Yorke J, Moosavi SH, Shuldham C, Jones PW. Quantification of dyspnoea using descriptors: development and initial testing of the Dyspnoea-12. Thorax. 2010;65(1):21–6.

26. Meek PM, Banzett R, Parsall MB, Gracely RH, Schwartzstein RM, Lansing R. Reliability and validity of the multidimensional dyspnea profile. Chest. 2012;141(6):1546–53.

27. Witek TJ, Jr., Mahler DA. Minimal important difference of the transition dyspnoea index in a multinational clinical trial. Eur Respir J. 2003;21(2):267–72.

28. Fletcher CM, Elmes PC, Fairbairn AS, Wood CH. The significance of respiratory symptoms and the diagnosis of chronic bronchitis in a working population. Br Med J. 1959;2(5147):257-66.

29. Zigmond AS, Snaith RP. The hospital anxiety and depression scale. Acta Psychiatr Scand. 1983;67(6):361–70.

30. Hibbard JH, Mahoney ER, Stockard J, Tusler M. Development and testing of a short form of the patient activation measure. Health Serv Res. 2005;40(6 Pt 1):1918-30.

31. Braun V, Clarke V. One size fits all? What counts as quality practice in (reflexive) thematic analysis? Qual Res Psychol. 2021;18(3):328–52.

32. Doe GE, Williams MT, Chantrell S, Steiner MC, Armstrong N, Hutchinson A, et al. Diagnostic delays for breathlessness in primary care: a qualitative study to investigate current care and inform future pathways. British Journal of General Practice. 2023:BJGP.2022.0475.

33. NHSEngland. Adult breathlessness pathway (pre-diagnosis): diagnostic pathway support tool 2023 [updated April 2023. Available from: https://www.england.nhs.uk/long-read/adult-breathlessness-pathway-pre-diagnosis-diagnostic-pathway-support-tool/.

34. Sunjaya AP, Homaira N, Corcoran K, Martin A, Berend N, Jenkins C. Assessment and diagnosis of chronic dyspnoea: a literature review. NPJ Prim Care Respir Med. 2022;32(1):10.

35. Doe G, Chantrell S, Williams M, Steiner MC, Armstrong N, Hutchinson A, et al. Breathless and awaiting diagnosis in UK lockdown for COVID-19…We’re stuck. NPJ Prim Care Respir Med. 2021;31(1):21.

36. Chowdhury SR, Chandra Das D, Sunna TC, Beyene J, Hossain A. Global and regional prevalence of multimorbidity in the adult population in community settings: a systematic review and meta-analysis. EClinicalMedicine. 2023;57:101860.

37. Panagioti M, Scott C, Blakemore A, Coventry PA. Overview of the prevalence, impact, and management of depression and anxiety in chronic obstructive pulmonary disease. Int J Chron Obstruct Pulmon Dis. 2014;9:1289–306.

38. Evans R, Pick A, Lardner R, Masey V, Smith N, Greenhalgh T. Breathing difficulties after covid-19: a guide for primary care. BMJ. 2023;381:e074937.

39. NICE. Asthma: diagnosis, monitoring and chronic asthma management. 2017 [Available from: https://www.nice.org.uk/guidance/ng80.

40. Olsson M, Björklund A, Sandberg J, et al. Factors most strongly associated with breathlessness in a population aged 50-64 years. ERJ Open 2024. Forthcoming

41. Ibrahim W, Natarajan S, Wilde M, Cordell R, Monks PS, Greening N, et al. A systematic review of the diagnostic accuracy of volatile organic compounds in airway diseases and their relation to markers of type-2 inflammation. ERJ Open Research. 2021;7(3):00030–2021.

42. Doe G, Taylor SJ, Topalovic M, Russell R, Evans RA, Maes J, et al. Spirometry services in England post-pandemic and the potential role of AI support software: a qualitative study of challenges and opportunities. British Journal of General Practice. 2023;73(737):e915–e23.

43. Moynihan R, Sanders S, Michaleff ZA, Scott AM, Clark J, To EJ, et al. Impact of COVID-19 pandemic on utilisation of healthcare services: a systematic review. BMJ Open. 2021;11(3):e045343.

44. Hull JH, Lloyd JK, Cooper BG. Lung function testing in the COVID-19 endemic. Lancet Respir Med. 2020;8(7):666–7.

45. White D, Hind D. Projection of participant recruitment to primary care research: a qualitative study. Trials. 2015;16(1):473.

46. NIHR. Equality, Diversity and Inclusion Strategy 2022-2027 2022 [Available from: https://www.nihr.ac.uk/documents/equality-diversity-and-inclusion-strategy-2022-2027/31295.

