## Supplement Information feasibility cRCT Breathe DEEP for "Investigating a structured diagnostic approach for chronic breathlessness in primary care: a mixed-methods feasibility cluster Randomised Controlled Trial"

### Figure 1. Investigations included in structured diagnostic pathway (Intervention)

| Investigation | Implications for diagnosis |
|---|---|
| **Body Mass Index (BMI)** | To diagnose obesity if >30kg/m$^2$ |
| **Electro cardiogram (ECG)** | To detect arrhythmias or diagnoses suggestive of ischaemic heart disease |
| **Chest X-ray** | To detect signs of heart failure or pulmonary pathology e.g. pleural effusion, interstitial changes |
| **Spirometry**<br>(If $FEV_1/FEV_6$ <0.75 perform full spirometry) | Obstructive pattern ($FEV_1$/FVC < 0.70) may suggest airway disease such as COPD or asthma<br>Restrictive pattern ($FEV_1$/FVC > 0.70 but $FEV_1$ and FVC < 80% predicted) may suggest conditions such as obesity, interstitial lung disease, chest wall abnormality or neuromuscular disease |
| **Full Blood Count (FBC)** | Haemoglobin to exclude anaemia<br>Eosinophil count if >0.3 x10^9/L can support a diagnosis of asthma |
| **NT-pro BNP profile** | To exclude heart failure |
| **Urea and electrolytes/ TSH** | To exclude metabolic causes |
| **Activity Questionnaire GPPAQ** | To assess physical activity levels |
| **PHQ4** | Screening questionnaires for symptoms of anxiety and depression |

*FEV1; Forced Expiratory Volume in 1 second, FEV6; Forced Expiratory Volume in 6 seconds, NT-pro BNP; N-terminal brain natriuretic peptide, TSH; Thyroid stimulating hormone, GPPAQ; General Practice Physical Activity Questionnaire, PHQ4; Patient Health Questionnaire 4 item.*

### Box 1.

| Feasibility measures |
|---|
| Number of patients recruited per week per GP practice population size |
| Number of participating GP practices verses the number approached |
| Time for GPs to screen for eligibility |
| Number of eligible patients who agree to be approached by the research team verses total number of eligible patients |
| Number and timing of investigations in the diagnostic pathway completed |
| Acceptability of the research visit to the participants |
| Data collected from Interviews regarding participant experience of the trial |

### Collection of Physical outcome measures

Physical outcome measures were collected for all participants attending a face-to-face research visit. Each participant's waist and hip circumference was measured and Body Mass Index (BMI, $kg/m^2$) was calculated by measuring the patient's height and weight. Body composition using bioelectrical impedance was measured to provide lean mass and body fat percentage. Exercise capacity was measured using the incremental shuttle walk test (ISWT) (3). Frailty was measured using Fried's Frailty definition (4), the Rockwood Frailty scale (5), the Timed up and Go and the Short Physical Performance battery (SPPB) (6). Physical Activity was assessed using the GT3x Actigraph device (7) and wrist worn GENEActiv device (8). All outcome measures and data collection methods are described in detail in the protocol paper (9).

### Spirometry

In the usual care group spirometry was not completed in 16 participants (13 not requested by clinician [6/13 no spirometry in primary care due to pandemic], 1 did not attend [DNA], 2 test contraindicated). In the intervention group spirometry was not completed in 11 participants (5 not requested by clinicians [3/5 no spirometry in primary care due to pandemic], 5 DNA, 1 contraindicated).

### Identify sources of data to plan the economic evaluation for a full trial

Primary care and hospital healthcare records were reviewed and data collected about number of GP consultations, referrals to secondary care, hospital outpatient appointments and hospital admissions (Table 1). An NHS Digital application to collect the healthcare utilisation data from Office for National Statistics (ONS) and Hospital Episodes Statistics (HES) was set up through the Data Access Request (DARS) portal; the appropriate data products to request were recorded and the information required has been mapped for a future, larger trial. We propose that this process would be required for a future trial and healthcare record review would not be feasible on a larger scale.

### Table 1. Healthcare Utilisation and Diagnoses related to breathlessness

| | **12 months** | |
|---|---|---|
| | Usual Care (n=23) | Intervention (n=25) |
| ***GP contacts*** | 4 (3-5) | 4 (2-4) |
| ***Referral:*** | | |
| *Respiratory* | 2 (9) | 3 (12) |
| *Cardiology* | 5 (22) | 4 (16) |
| *Breathlessness Service* | 1 (4.5) | 3 (12) |
| *Other* | 2 (9) | 0 |
| ***Coded Diagnosis*** | 6 (26) | 11 (44) |
| ***Outpatient appointments*** | | |
| *0* | 17 (73.1) | 15 (60) |
| *1* | 0 | 3 (12) |
| *2* | 4 (17.2) | 4 (16) |
| *3* | 1 (4.3) | 1 (4) |
| *4* | 0 | 2 (8) |
| *5* | 0 | 0 |
| *6* | 1 (4.3) | 0 |
| ***Hospital admissions*** | 3 (12) | 1 (14) |

*Data presented as frequency (%) or Median (IQR)*

Figure 2. Monthly breakdown of eligible patients from GP practices

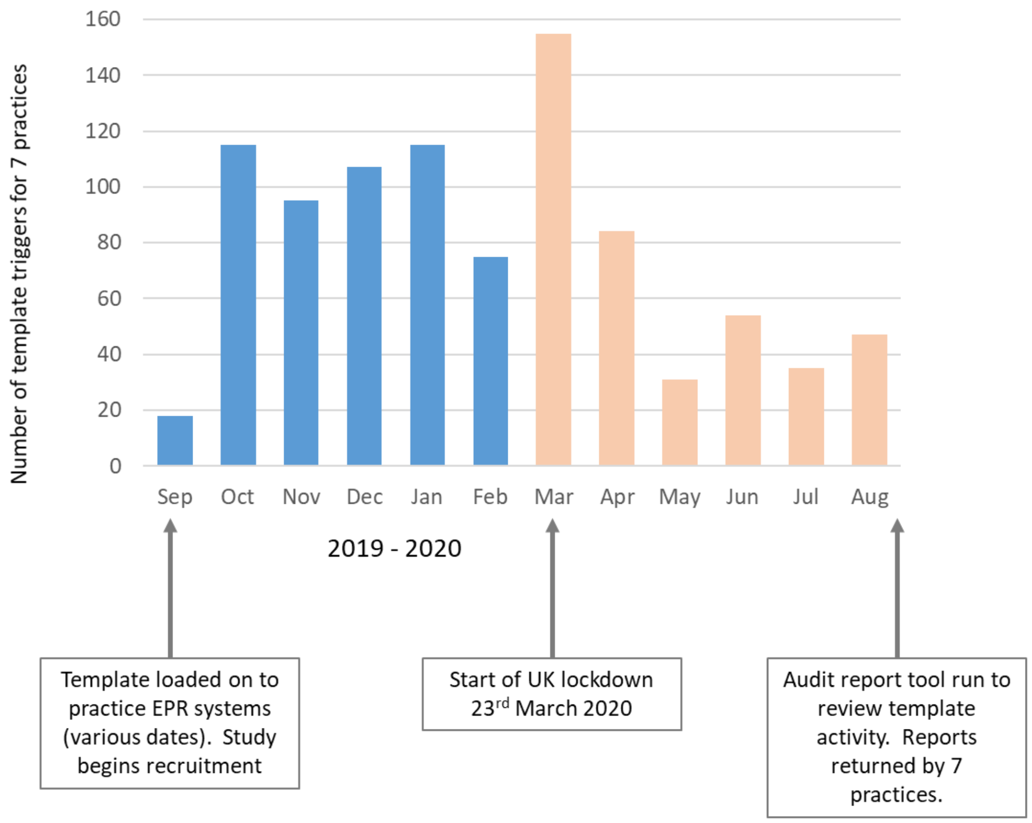


An audit tool on the electronic patient record (EPR) system was used to measure the usage of the electronic template recruitment aid, establishing the proportion of patients willing to be approached by the study team verses those eligible. Data available from 7 of the 10 GP practices showed between 6 and 19% of patients identified as eligible from practices were sent to the study team. The proportion of remaining patients were either identified as not eligible by the GP, declined to be contacted by the study team or had the template closed down without further action. The audit tool also measured the number of patients possibly eligible for the trial (presenting with breathlessness) for the period September 2019 to February 2020 (pre pandemic, n = 525) compared with March to September 2020 (n = 406), to assess the impact of the pandemic on patients presenting with breathlessness in primary care.

### Feasibility of data collection

Thirty-two participants completed in person research visits at baseline as per the original study design, 16 completed their consent and research visit by phone. Of those who attended an in-person research visit, 23/32 (72%) completed the exercise capacity ISWT. Reasons for non-completion was elevated blood pressure beyond the acceptable limits for the walk test and mobility limited by pain. Forty-seven of the forty-eight participants recruited returned the PROM questionnaires at baseline. The number of participants able to complete each outcome measurement at baseline is included in Table 2.

A large proportion of the secondary outcome measures assessing physical function were missing from the 12 month follow up due to the change in study protocol which moved to remote data collection as a result of the pandemic (supplementary Table 2). Due to this incomplete data set of physical measures, only PROMs results are presented for comparison at six and twelve months to baseline (Figure 4). No serious adverse events (SAEs) were recorded for this study.

Ease of use of PROM questionnaires was assessed by missing data and support required from research team with follow up questionnaires. From 82 questionnaire follow ups (41 at six and twelve months), the CHQ required help from one participant compared with a range of 3 to 10 instances for other questionnaires.

#### Table 2. Physical Outcome measures missing data at 12 months

| **Outcome measure** | **Not completed n** | **Usual care n (%)** | **Female n (%)** |
|---|---|---|---|
| *ISWT* | 30 | 15 (50) | 21 (70) |
| *QMVC* | 25 | 14 (56) | 18 (72) |
| *Handgrip* | 24 | 14 (58) | 18 (72) |
| *SPPB* | 24 | 14 (58) | 18 (72) |
| *TUG* | 23 | 14 (61) | 18 (72) |
| *4MGS* | 24 | 14 (58) | 18 (72) |
| *Bioelectrical impedance* | 25 | 15 (60) | 18 (72) |

44 Participants completed a follow up visit at 12 months; 20 in person and 24 by phone, 41 returned questionnaires.

### Table 3. Number and proportion of investigations completed

| | 3 months | | 12 months | |
|---|---|---|---|---|
| **Investigation** | Usual Care n = 23 | Intervention n = 25 | Usual Care n = 23 | Intervention n = 25 |
| ***Body Mass Index (BMI)*** | 12 (52) | 14 (56) | 16 (70) | 21 (84) |
| ***Chest X-ray*** | 15 (65) | 23 (92) | 20 (87) | 25 (100) |
| ***Electrocardiogram (ECG)*** | 10 (44) | 19 (76) | 15 (65) | 20 (80) |
| ***Spirometry*** | 5 (22) | 11 (44) | 7 (30) | 14 (56) |
| ***Full Blood Count (FBC)*** | 18 (78) | 21 (84) | 20 (87) | 23 (92) |
| ***NT-proBNP*** | 13 (57) | 20 (80) | 16 (70) | 21 (84) |
| ***Urea and Electrolytes*** | 17 (74) | 22 (88) | 20 (87) | 23 (92) |
| ***Thyroid Stimulating Hormone*** | 13 (57) | 19 (76) | 17 (74) | 20 (80) |
| ***PHQ-4*** | 1 (4) | 20 (80) | 2 (8) | 21 (84) |
| ***GPPAQ*** | 1 (4) | 18 (72) | 1 (4) | 20 (80) |
| ***All above*** | 0 | 4 (16) | 0 | 8 (32) |
| ***All above minus Spirometry*** | 0 | 5 (20) | 0 | 11 (44) |

*PHQ-4 = Patient health questionnaire 4 item, GPPAQ = GP Physical Activity Questionnaire.*

*Data presented as Mean (SD), Median (IQR) or frequency (%)*

### Figure 3. Coded Diagnostic labels at 12 months

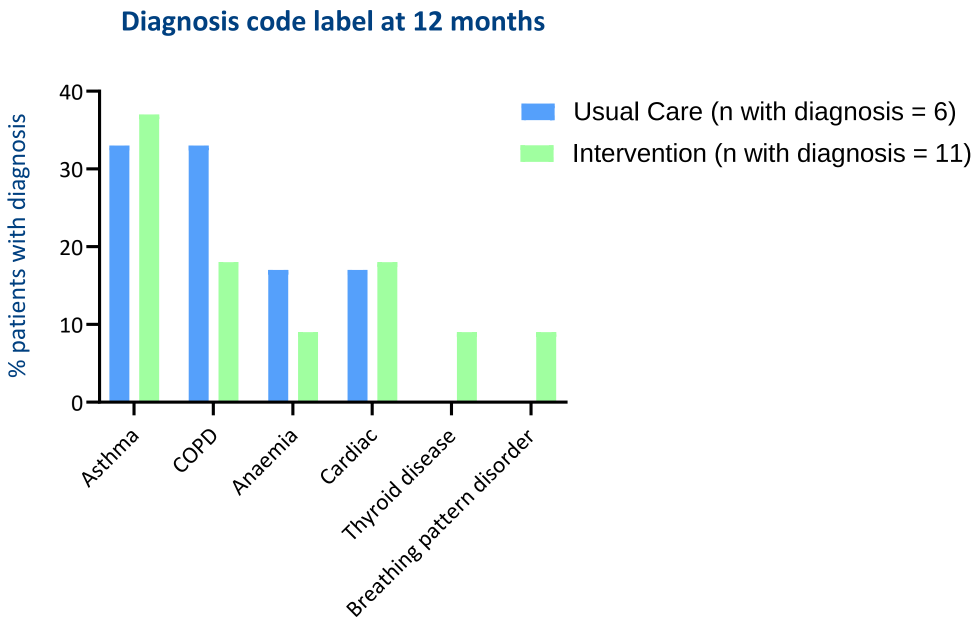


*Coded diagnoses identified through healthcare record review at 12 months, proportion of participants in each group with these diagnoses. 11 participants had a coded diagnosis at 12 months in the Intervention group, 6 participants had a coded diagnosis in Usual care.*

### Box 2

**Intraclass Correlation Coefficient**

*Calculation using the Dyspnoea domain of the CHQ*

Using a SD of 1.61 and minimal clinically important difference (MCID) for the CHQ of 0.5 with loss to follow up of 15%, the sample required for 80% power at the 5% significance level before inflating for clustering is 386 participants (159 per arm); 328 overall before accounting for loss to follow up.

With an estimate of the ICC of 0.1, derived from the participant data in this study from the CHQ Dyspnoea domain, the inflation factor is around 1.69 (based on mean cluster size of 7, minimum 2, maximum 12 after lose to follow up; ICC=0.1) so the total sample required would be 660 randomised (330 per arm from 40 clusters); 560 overall before accounting for loss to follow up.

*Calculation using Mastery domain of the CHQ*

Using a SD of 1.46 and minimal clinically important difference (MCID) for the CHQ of 0.5 with loss to follow up of 15%, the sample required for 80% power at the 5% significance level before inflating for clustering is 318 participants (159 per arm); 270 overall before accounting for loss to follow up.

With an estimate of the ICC of 0.05, derived from the participant data in this study from the CHQ Mastery domain, the inflation factor is around 1.35 (based on mean cluster size of 7, minimum 2, maximum 12 after lose to follow up; ICC=0.05) so the total sample required would be 430 randomised (215 per arm from 26 clusters); 364 overall before accounting for loss to follow up.

The recruitment and retention achieved in this feasibility study has allowed estimation of an ICC to aid future realistic sample and cluster sizes for a larger trial which is important for achieving successful research outcomes. We propose a possible primary outcome measure for a future trial could be number and time to diagnosis or a measure of symptom burden or health related quality of life. PROMs collected in this trial allowed an indication of which might be a suitable choice. We propose that future outcome measures aligned to health-related quality of life could be the CHQ or EQ5D-5L. The CHQ measures symptom burden (Dyspnoea domain) and mastery/control over symptoms (Mastery domain). Economic analysis would be required for a future trial and the EQ5D is therefore also a well-validated and widely used tool to consider.(1, 2)

### Table 4. All Physical Outcome measures collected at baseline and 12 months

| | All Participants | | | | Usual Care | | | | Intervention | | | |
|---|---|---|---|---|---|---|---|---|---|---|---|---|
| | n | Baseline | n | 12 months | n | Baseline | n | 12 months | n | Baseline | n | 12 months |
| ***ISWT (m)*** | 23 | 348 (196) | 14 | 451 (158) | 9 | 426 (217) | 6 | 522(161) | 14 | 299 (170) | 8 | 399 (143) |
| *SpO2 post-ISWT (%)* | | 92 (4) | | 93 (3) | | 93 (4) | | 95(2) | | 92 (4) | | 92 (4) |
| *Peak HR (bpm)* | | 92 (18) | | 102 (13) | | 103 (17) | | 112 (12) | | 85 (15) | | 94 (7) |
| *Peak BORG* | | 3.0 (3.0-4.0) | | 3.5 (2.7-4.8) | | 3.5 (2.5-4.0) | | 4.0 (3.5-4.3) | | 3.5 (3.0-4.0) | | 3.0 (2.3-3.8) |
| *Reason for terminating test:* | | | | | | | | | | | | |
| *Breathlessness* | | | | 10 (71) | | | | 4 (67) | | | | 6 (75) |
| *Pain* | | | | 1 (7) | | | | 0 | | | | 1 (12.5) |
| *Breathless/Leg fatigue* | | | | 2(15) | | | | 1 (16.5) | | | | 1 (12.5) |
| *Other* | | | | 1 (7) | | | | 1 (16.5) | | | | 0 |
| ***QMVC (kg)*** | 30 | 28.7 (15.1) | 19 | 30.5 (14.6) | 10 | 28.0 (14.8) | 7 | 35.5 (16.4) | 20 | 29.0 (15.6) | 12 | 27.5 (13.2) |
| ***Handgrip (kg)*** | 31 | 30.4 (11.8) | 20 | 32.8 (13.5) | 11 | 29.5 (4.3) | 7 | 38.5 (15.6) | 20 | 30.8 (2.4) | 13 | 29.6 (11.8) |
| ***SPPB (score)*** | 31 | 9.0 (7.0-11.0) | 20 | 10.5 (9.0-12.0) | 11 | 8.0 (7.0-11.0) | 7 | 11 (10.0-12.0) | 20 | 9.0 (7.0-10.8) | 13 | 9.0 (8.0-11.5) |
| ***TUG (seconds)*** | 30 | 8.9 (7.1-11.3) | 20 | 8.2 (6.6-8.9 | 11 | 9.7 (6.4 -17.9) | 7 | 6.9 (6.3-8.2) | 19 | 8.9 (7.1-10.8) | 13 | 8.4 (7.4-9.7) |
| ***4MGS (seconds)*** | 31 | 4.0 (3.5-5.2) | 20 | 4.0 (3.5-4.6) | 11 | 3.9 (3.6-5.6) | 7 | 3.6 (3.1-4.1) | 20 | 4.1 (3.4-5.1) | 13 | 4.3 (3.9-5.1) |
| ***Fried's Frailty:*** | | | | | | | | | | | | |
| *Robust* | 29 | 7 (24) | 17 | 5 (29) | 11 | 4 (36) | 7 | 4 (57) | 18 | 3 (17) | 10 | 1 (10) |
| *Pre Frail* | | 20 (69) | | 12 (71) | | 5 (46) | | 3 (43) | | 15 (83) | | 9 (90) |
| *Frail* | | 2 (7) | | 0 | | 2 (18) | | 0 | | 0 (0) | | 0 |
| ***Body fat (kg)*** | 32 | 34.1 (12.0) | 19 | 33.7 (13.3) | 12 | 31.5 (10.0) | 6 | 26.0 (6.7) | 20 | 35.6 (13.0) | 13 | 37.2 (14.3) |
| ***Body fat (%)*** | 32 | 39.4 (9.0) | 19 | 38.1 (8.5) | 12 | 38.6 (9.9) | 6 | 31.5 (7.3) | 20 | 39.9 (8.7) | 13 | 41.2 (7.4) |
| ***Physical Activity:*** | | | | | | | | | | | | |
| *Step count (Actigraph)* | 32 | 5011 (2560) | 27 | 4753 (2853) | 11 | 5041 (3090) | 13 | 4772 (3034) | 21 | 4996 (2320) | 14 | 4735 (2790) |
| *Overall Activity (mg)* | 29 | 21.8 (7.7) | 26 | 20.2 (6.1) | 12 | 21.0 (6.0) | 13 | 18.7 (5.8) | 17 | 19.5 (5.9) | 13 | 21.7 (6.2) |
| **Time Inactive (mins)* | 29 | 660 (113) | 26 | 674.7 (63.5) | 12 | 649 (93) | 13 | 670.2 (65.8) | 17 | 668 (128) | 13 | 679.1 (63.5) |

Data is presented as Mean (SD), frequency (%) or Median (IQR). *HR = heart rate, QMVC = quadriceps maximal voluntary contraction, SPPB = short performance physical battery, TUG = timed up and go, 4MGS = 4 metre gait speed*. *Time Inactive defined as time not moving including standing and therefore different to sedentary time (sitting, lying or reclining).

### Figure 4 Patient Reported Outcome Measures

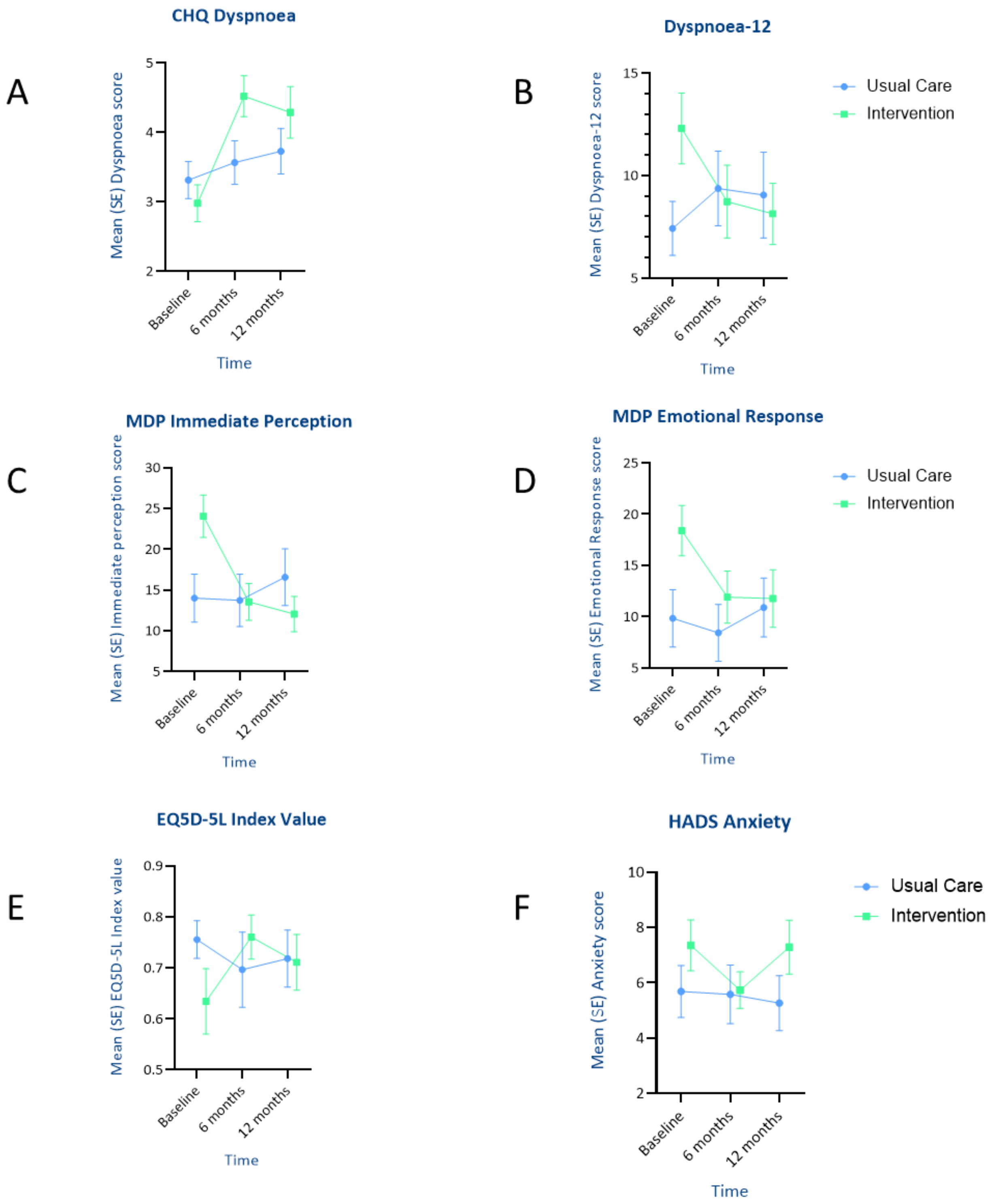


*Mean score and Standard Error (SE) for PROMs at baseline, 6 and 12 months. A. Chronic Heart Questionnaire (CHQ) Dyspnoea domain; B. Dyspnoea-12 questionnaire; C. Multidimensional Dyspnoea Profile (MDP) Immediate perception; D. Multidimensional Dyspnoea Profile (MDP) Emotional Response; E. EuroQoL 5 Dimension 5 level (EQ5D-5L) Index value score; F. Hospital Anxiety and Depression (HADs) questionnaire Anxiety score*

### Table 5. Baseline and Transition Dyspnoea Index

| | All Participants | | Usual Care | | Intervention | |
|---|---|---|---|---|---|---|
| | n | | n | | n | |
| *BDI focal score* | 48 | 6.4 (2.1) | 23 | 6.2 (2.3) | 25 | 6.7 (2.0) |
| *TDI focal score 6 months* | 39 | 1.1 (3.7) | 18 | 0.3 (3.2) | 21 | 1.9 (3.9) |
| *TDI focal score 12 months* | 44 | 0.6 (4.3) | 21 | -0.9 (3.7) | 23 | 2.0 (4.5) |

Data is presented as Mean (SD), *BDI = Baseline Dyspnoea Index (focal score ranges from 0-12 with 0 representing worse impairment), TDI = Transition Dyspnoea Index (focal score ranges from -9 to +9 with -scores being a deterioration and +scores being an improvement).*

### References


1. Devlin NJ, Brooks R. EQ-5D and the EuroQol Group: Past, Present and Future. Appl Health Econ Health Policy. 2017;15(2):127-37.
2. (5Q5D5L). E. Euroqol (5Q5D-5L) 2016 [Available from: www.euroqol.org/eq-5d-products/eq-5d-5l.html .
3. Singh SJ, Morgan MD, Scott S, Walters D, Hardman AE. Development of a shuttle walking test of disability in patients with chronic airways obstruction. Thorax. 1992;47(12):1019-24.
4. Fried LP, Tangen CM, Walston J, Newman AB, Hirsch C, Gottdiener J, et al. Frailty in older adults: evidence for a phenotype. J Gerontol A Biol Sci Med Sci. 2001;56(3):M146-56.
5. Rockwood K, Song X, MacKnight C, Bergman H, Hogan DB, McDowell I, et al. A global clinical measure of fitness and frailty in elderly people. CMAJ. 2005;173(5):489-95.
6. Guralnik JM, Simonsick EM, Ferrucci L, Glynn RJ, Berkman LF, Blazer DG, et al. A short physical performance battery assessing lower extremity function: association with self-reported disability and prediction of mortality and nursing home admission. J Gerontol. 1994;49(2):M85-94.
7. Actigraph.
8. GeneActiv. [Available from: https://www.activinsights.com/actigraphy/geneactiv-original/
9. Doe G, Clanchy J, Wathall S, Chantrell S, Edwards S, Baxter N, et al. Feasibility study of a multicentre cluster randomised control trial to investigate the clinical and cost-effectiveness of a structured diagnostic pathway in primary care for chronic breathlessness: protocol paper. BMJ Open. 2021;11(11):e057362.
